# Mode of inheritance needs to be accounted for in interpreting genotype-phenotype links in monogenic disorders

**DOI:** 10.1101/2023.09.26.23296051

**Authors:** Antoni Riera-Escamilla, Corrine Kolka Welt, Maris Laan

**Author notes:** **Matters Arising from:** Ke H, Tang S, Guo T, Hou D, Jiao X, Li S, Luo W, Xu B, Zhao S, Li G, Zhang X, Xu S, Wang L, Wu Y, Wang J, Zhang F, Qin Y, Jin L, Chen ZJ. Landscape of pathogenic mutations in premature ovarian insufficiency. *Nat Med* 29, 483–492 (2023). doi: 10.1038/s41591-022-02194-3.

## Abstract

**Introduction:** A recently published study by Ke *et al*. utilized whole exome sequencing (WES) to screen genetic variants contributing to premature ovarian insufficiency (POI) in a large cohort of 1,030 patients from China (doi: 10.1038/s41591-022-02194-3). The authors reported that 285 likely pathogenic (LP) and pathogenic (P) variants identified in 79 genes contributed to POI in 242 study subjects, representing 23.5% of the cohort. The majority, 191 patients (∼79%), carried monoallelic (heterozygous) variants.

**Objective:** We re-analyzed the contribution of reported genotypes considering the inheritance mode of POI and other inherited conditions linked to 79 genes with reported findings by Ke *et al*.

**Methods:** The disease inheritance modes linked to targeted genes were retrieved from publicly available databases (OMIM, Genomic England PanelApp, PubMed, DOMINO, gnomAD). Genotypes of 242 cases reported by Ke *et al.* were assessed in the context of known inheritance mode(s) of disorders linked to respective genes.

**Results:** Most, 48 of 79 genes were classified as recessive, whereas only 13 genes were dominant. Insufficient data was available for 18 genes to conclusively determine their inheritance mode. Nearly half of 242 cases reported by Ke *et al*., 119 women (∼49%), carried heterozygous variants in known autosomal recessive genes and therefore these variants are not contributing to their POI phenotype. Only 68 of women (6.6%) carried biallelic variants in either recessive or dominant genes or monoallelic variants in dominant genes, hence contributing to the diagnostic yield. This is ∼3.5-fold lower than 23.5% claimed in Ke *et al*. Additional 56 women (5.4%) were reported monoallelic variants in genes with insufficient data to determine the inheritance mode or multiple heterozygous variants in >1 recessive gene, whereby oligogenic contribution to POI cannot be excluded. But when even including these cases, the maximum estimated contributing yield is ∼12%, two times lower than claimed.

**Conclusion:** Using WES to screen monogenic causes of POI as part of the diagnostic pipeline will improve patient management strategies, but overestimated diagnostic yield in genetic research can create unrealistic expectations in the POI clinical community, typically non-specialist in genetics.

---

We have read with interest the recently published paper from Ke *et al*. wherein they utilized whole exome sequencing (WES) to screen genetic variants contributing to premature ovarian insufficiency (POI) in a large cohort of 1,030 patients from China^1^. POI is defined as the loss of normal ovarian function before the age of 40 years, affecting ∼1% of women^2^. The etiology of POI is broad, including well-established genetic factors explaining 20-25% of cases, such as X-monosomy (45,X0; Turner syndrome) or a premutation in the *FMR1* gene^2^. Despite multiple efforts, ∼70% cases remain unresolved. Availability of whole exome (WES) sequencing has promoted the search of monogenic causes of POI. So far, most studies have used WES to identify monogenic causes of POI in case studies and/or cohorts of families with the condition and there are limited data on the yield of WES in larger clinical cohorts of unrelated subjects^3–6^.

As the main outcome of the study, Ke *et al*. reported that the rate of contribution to POI by genetic variations uncovered from the WES data reached 23.5%. We read the study with interest to understand in detail its design in achieving this high yield. First, Ke *et al*. screened for pathogenic or likely pathogenic (P/LP) variants on 95 well-established POI-causative genes reported in the OMIM database (https://www.omim.org) or scientific literature. For 193 patients, 195 distinct variants were identified in 59 known POI genes. To uncover novel POI candidate genes, the authors tested the burden of rare and potentially pathogenic variants in pre-selected 646 genes in the POI cohort of 1,030 women compared to 5,000 population controls from the HuaBiao project (men and women, aged 16–83 y recruited across China)^7^. This analysis identified 20 novel POI candidate genes (**Fig. 1a**) with 59 loss-of-function variants detected in 61 cases, out of which 49 did not have findings in the known POI gene set.

**Figure 1.**
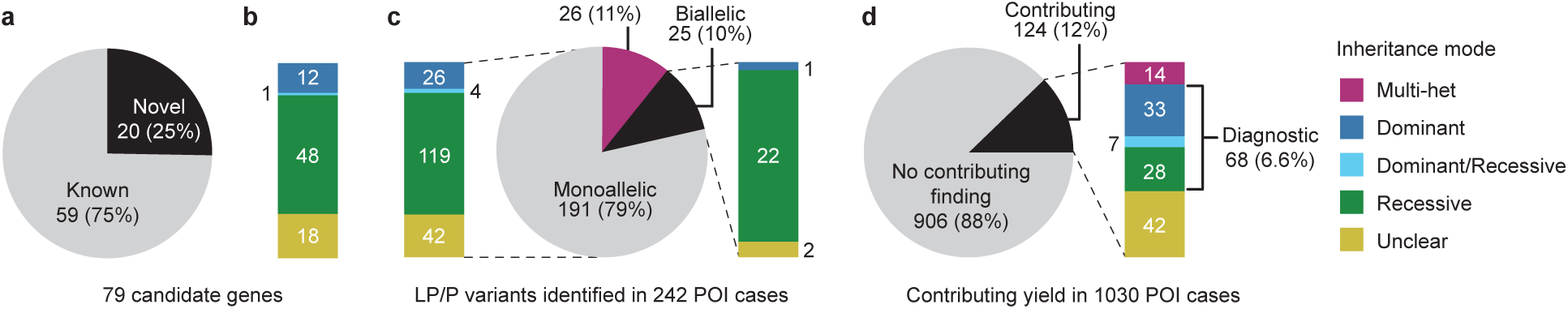
Assessment of the gene inheritance mode and the contribution of genotypes reported by Ke et al. to premature ovarian insufficiency (POI). **(a)** 59 known and 20 novel POI candidate genes reported Ke *et al*. with likely pathogenic/pathogenic (LP/P) variants identified in 242 of 1,030 analyzed patients. **(b)** Candidate genes categorized by known inheritance mode of linked diseases derived from public databases and/or literature. **(c)** Classification of genotypes in 242 POI cases by Ke *et al*. (circle) and reported mono-and biallelic variants categorized according to the inheritance mode linked to the affected genes (columns); ‘Multi-het’ refers to cases with LP/P variants in >1 gene, and ‘Unclear’ indicates genes with insufficient data of inheritance mode. **(d)** Maximum potential contributing and diagnostic yield of genetic findings in 1,030 POI cases after considering reported genotypes in the context of gene inheritance modes; contributing yield includes all findings by Ke *et al*. except cases with monoallelic variants in a single recessive gene, whereas diagnostic yield includes only clinically valid findings (biallelic variants in any candidate gene and monoallelic variants in dominant or dominant/recessive genes).

In total, the authors reported 285 P/LP variants in 79 genes contributing to the POI phenotype in 242 of 1,030 study subjects (23.5%). The majority, 191 patients (∼79%), carried monoallelic (heterozygous) variants. For dominant genes, a heterozygous pathogenic change is usually sufficient to cause the disease phenotype. Conversely, carriers of a monogenic pathogenic allele in a recessive gene are asymptomatic on most occasions and only biallelic mutations (homozygous or compound heterozygous) have a direct causal link to the phenotype. However, assessment of the primary data of the Ke *et al*. study revealed that the mode of inheritance has not been always properly accounted for in interpreting the pathogenicity of monoallelic variants and their confident contribution to POI. The authors appeared to have deviated from the ACMG standards for classification of genotype pathogenicity, and conflated the concepts of “contribution”, “contribution yield” and “causality,” potentially leading to inflated interpretation of the data.

To clarify the genetic contributing yield and the causality (genetic diagnosis) of the variants identified in the gene panel to POI, we reassessed the pertinent results of the study. First, we categorized the involved 79 genes according to the inheritance mode of linked disorders (**Fig. 1b**, **Supplementary Table 1**). This categorization drew upon publicly available expert evaluations sourced from OMIM (https://www.omim.org) and the Genomic England PanelApp (https://panelapp.genomicsengland.co.uk), haploinsufficiency data in the gnomAD (https://gnomad.broadinstitute.org) and DOMINO platforms (https://domino.iob.ch/about.php), and scientific literature. Approximately 61% (48 of 79) genes were confidently classified as recessive genes, whereby only biallelic pathogenic variants contribute to the phenotype. Only 13 genes were linked to dominant disorders. The available data for 18 genes (in this commentary referred as ‘Unclear’) was insufficient to conclusively determine their specific inheritance mode.

Next, we assessed the genotypes of the reported 242 POI patients according to the inheritance mode of the respective gene. Only 30 of 191 patients (15.7%) with monoallelic variants reported by Ke *et al*. had findings in established dominant genes or loci linked to both dominant and recessive disorders (**Fig. 1c**, **Supplementary Table 2**). Additional seven of 26 cases categorized by the authors as ‘Multi-het’ (referring to LP/P variants identified in more than one gene) had monoallelic findings in at least one dominant or dominant/recessive gene. However, the majority of monoallelic LP/P variants reported by Ke *et al* (119 of 191 cases; 62.3%) were found in genes linked to recessive disorders. An example of a well-established autosomal-recessive (AR) gene is *FSHR*, the first discovered POI locus^8^. Other examples of recessive POI genes with monoallelic findings by Ke et al. are *SPIDR*^9,10^, *MSH4*^11^*, EIF2B2*^12^, and *MCM9*^13^. Conducted family-based studies have identified no POI phenotype in the heterozygous mothers (carriers of damaging variants) of affected homozygous daughters. Likewise, carriers of monoallelic *BRCA2* pathogenic variants do not have reduced ovarian reserve and only biallelic disease-causing variants have been confidently linked to POI in previous studies^14^.

In a recent study of 102,502 UK biobank participating females with age at menopause >40 years, high number of carriers of heterozygous high-confidence protein-truncating variants were reported in the *FSHR* (n=72)*, SPIDR* (n=150)*, EIF2B2 (n=83), MCM9* (n=65) and *BRCA2* (n=317) genes^15^ (**Supplementary Table 1**). These data are in line with the evidence from numerous family-based studies and further emphasize a lack of major phenotypic effect of loss-of-function monoallelic variants in recessive genes linked to POI.

As a further note, Ke et al. has commented that ‘pLI could serve as a rough guide for prioritizing new POI genes’. However, pLI (the probability of being loss-of-function intolerant) scores were not always considered in prioritizing novel genes or in the analysis of pathogenicity of genotypes. Overall, 16 of 20 novel claimed POI genes had pLI 0 – 0.08, indicating tolerance to monoallelic pathogenic variants. Only four novel candidate genes (*CPEB1, PRDM1, BMP6, LGR4*) showed confident signs of haploinsufficiency (pLI = 0.49 – 1), supported by other lines of evidence.

After reassessment of genotypes in 242 POI cases highlighted by Ke *et al*., we estimated the maximum potential contributing yield of the reported variants to be ∼12% (124 of 1,030 patients; **Fig. 1d**, **Supplementary Table 2**. This non-stringent estimate also includes 56 women (5.4%) with monoallelic variants in genes with insufficient data for the inheritance mode (‘Unclear’) or identified in more than one recessive gene (‘Multi-het’). The actual clinical diagnostic yield was estimated 6.6% (68 of 1030 POI cases), when accounting for the known inheritance mode of involved genes. This is ∼3.5-fold lower than 23.5% claimed in the publication by Ke *et al*.

We believe that using WES or targeted gene panel to screen monogenic causes of POI as part of the diagnostic pipeline will potentially improve patient management strategies, as well as clinical counselling and decision making. However, only pathogenic genotypes with confident causal links to the condition have clinical significance and value. Incorrect interpretation of the contribution of genetic findings and significant overestimation of their yield will lead to biased expectations in the POI clinical community, who typically does not have specialist expertise in medical genetics.

## Data availability statement

All relevant data is provided in the article and its accompanying supplementary materials.

## Data Availability

All data produced in the present work are contained in the manuscript.

---

**Supplementary Table 1.**
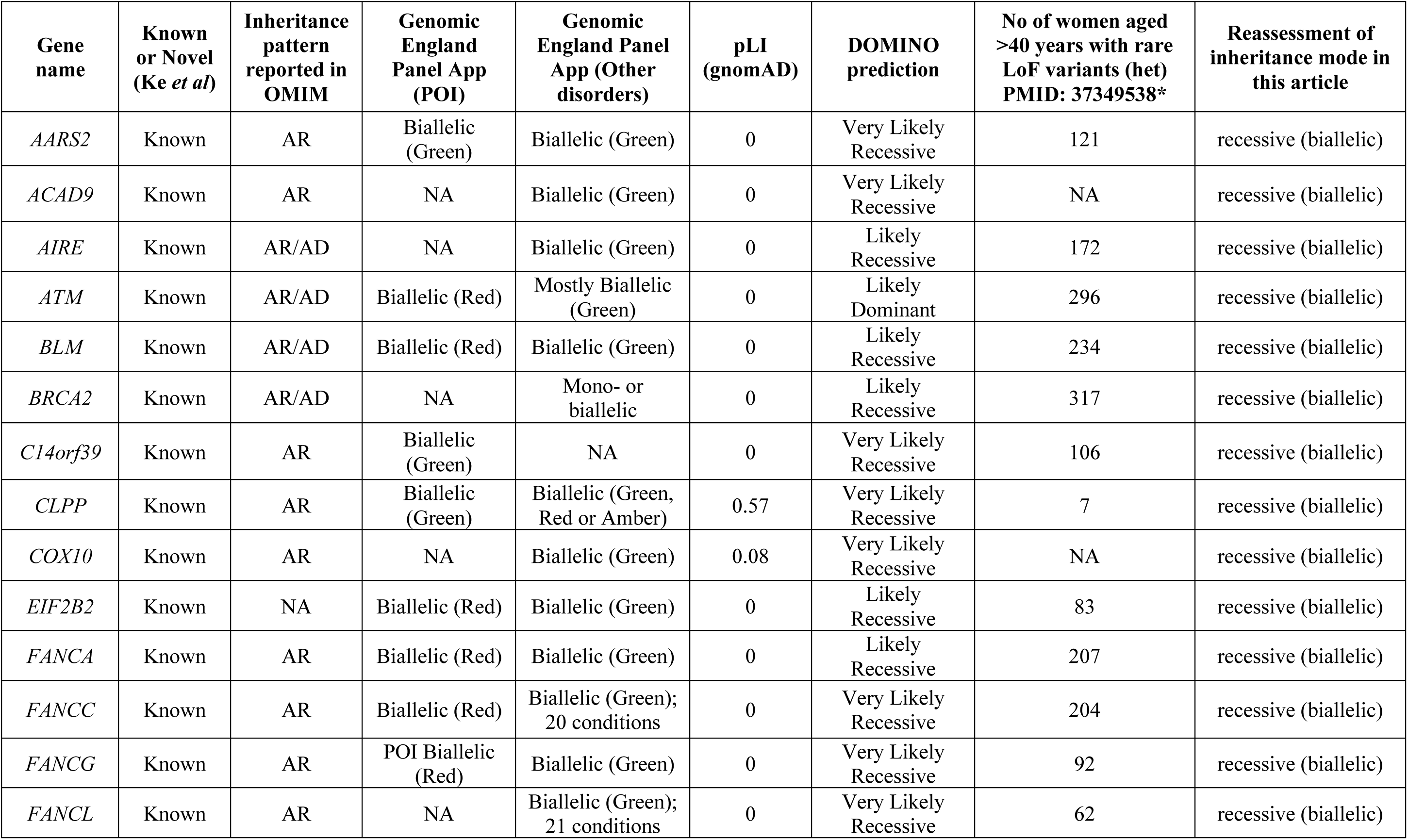
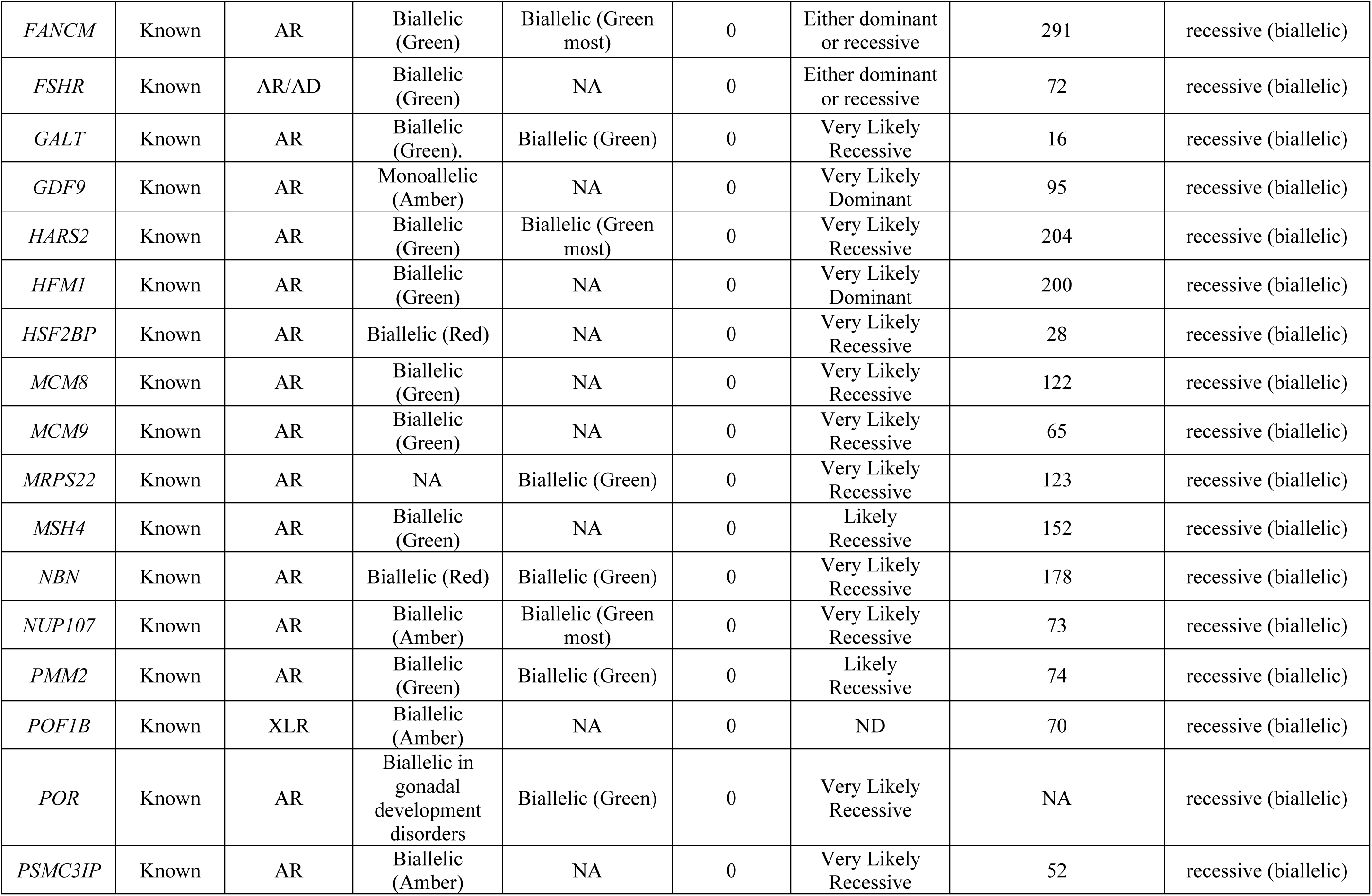
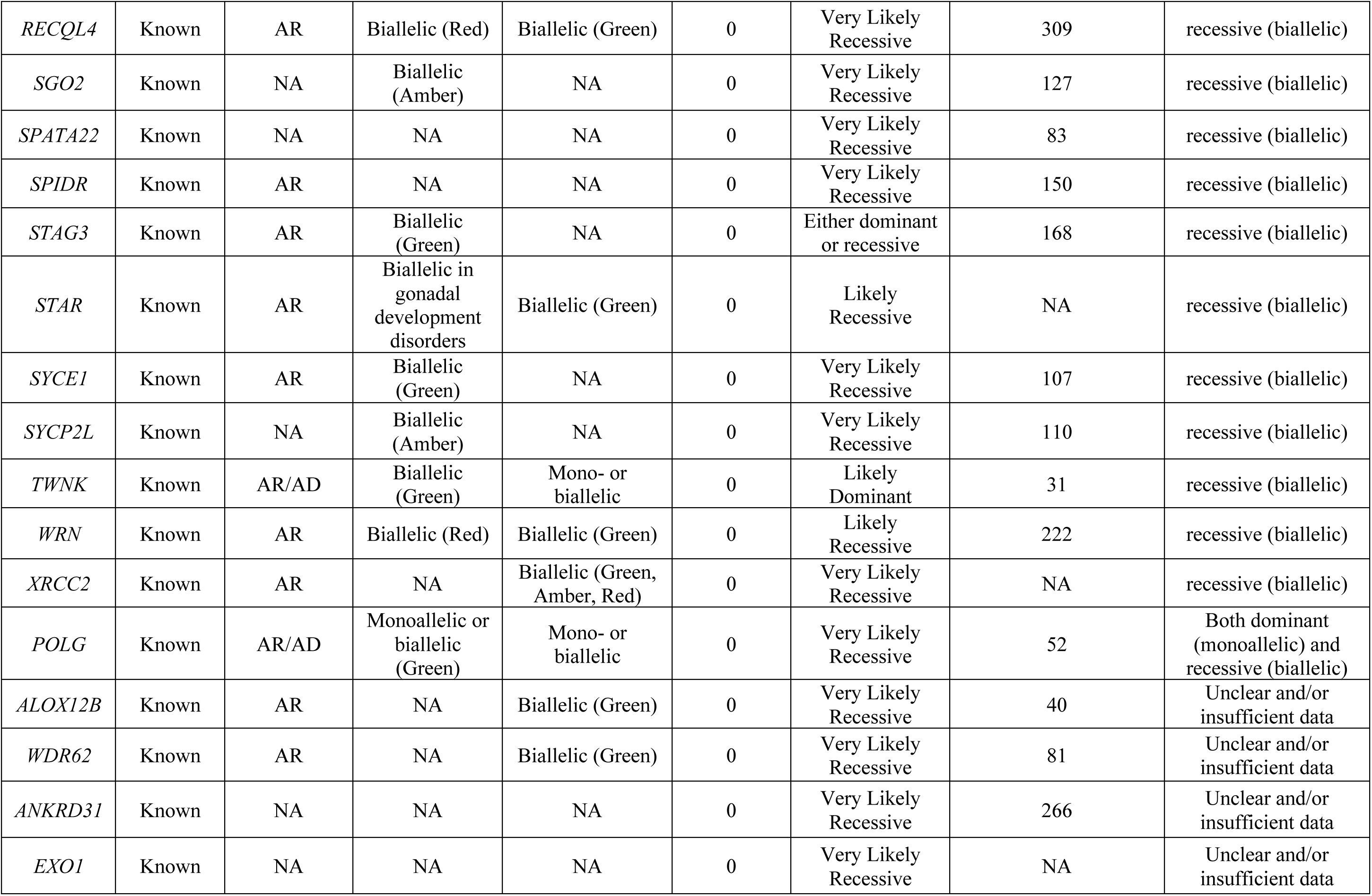
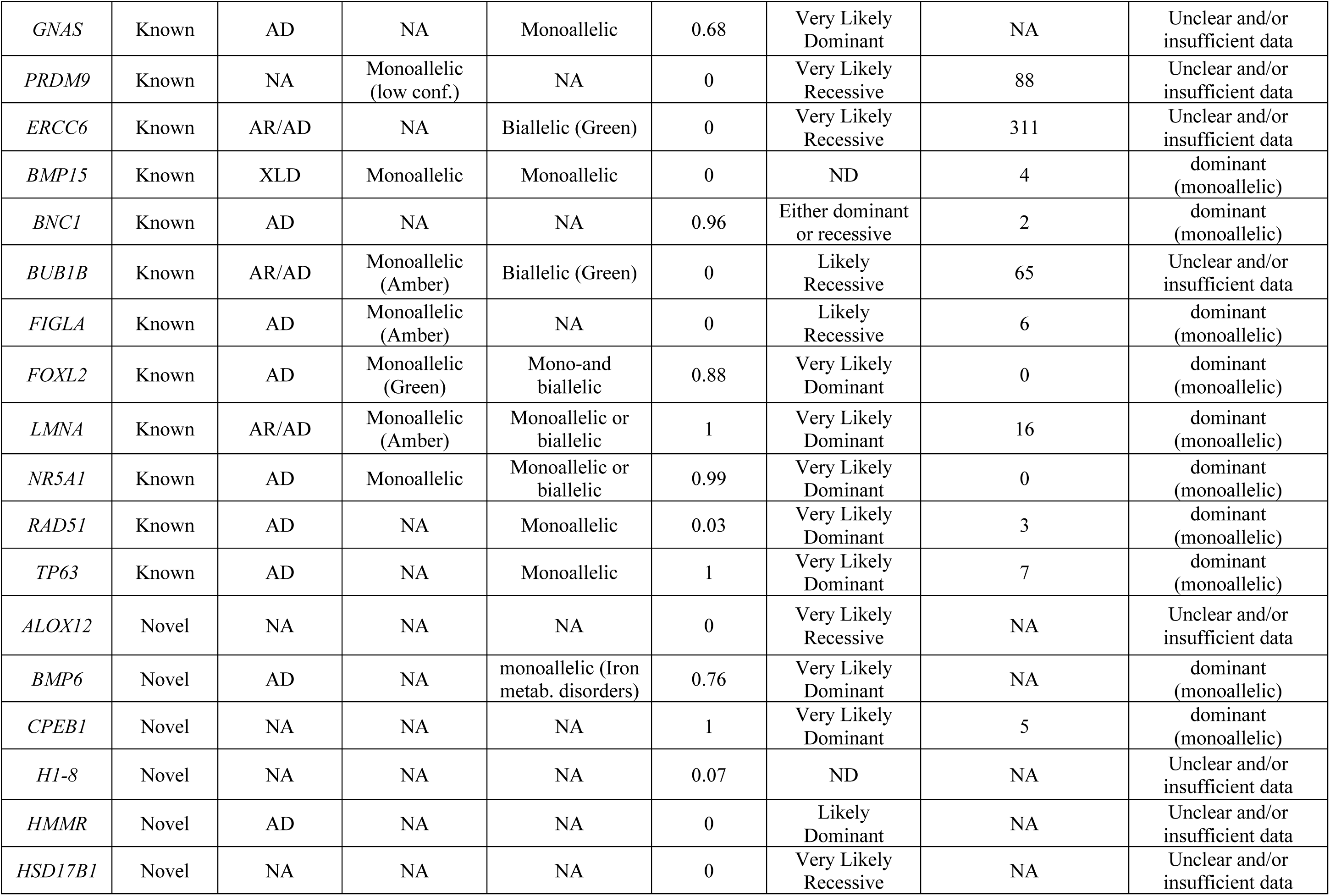
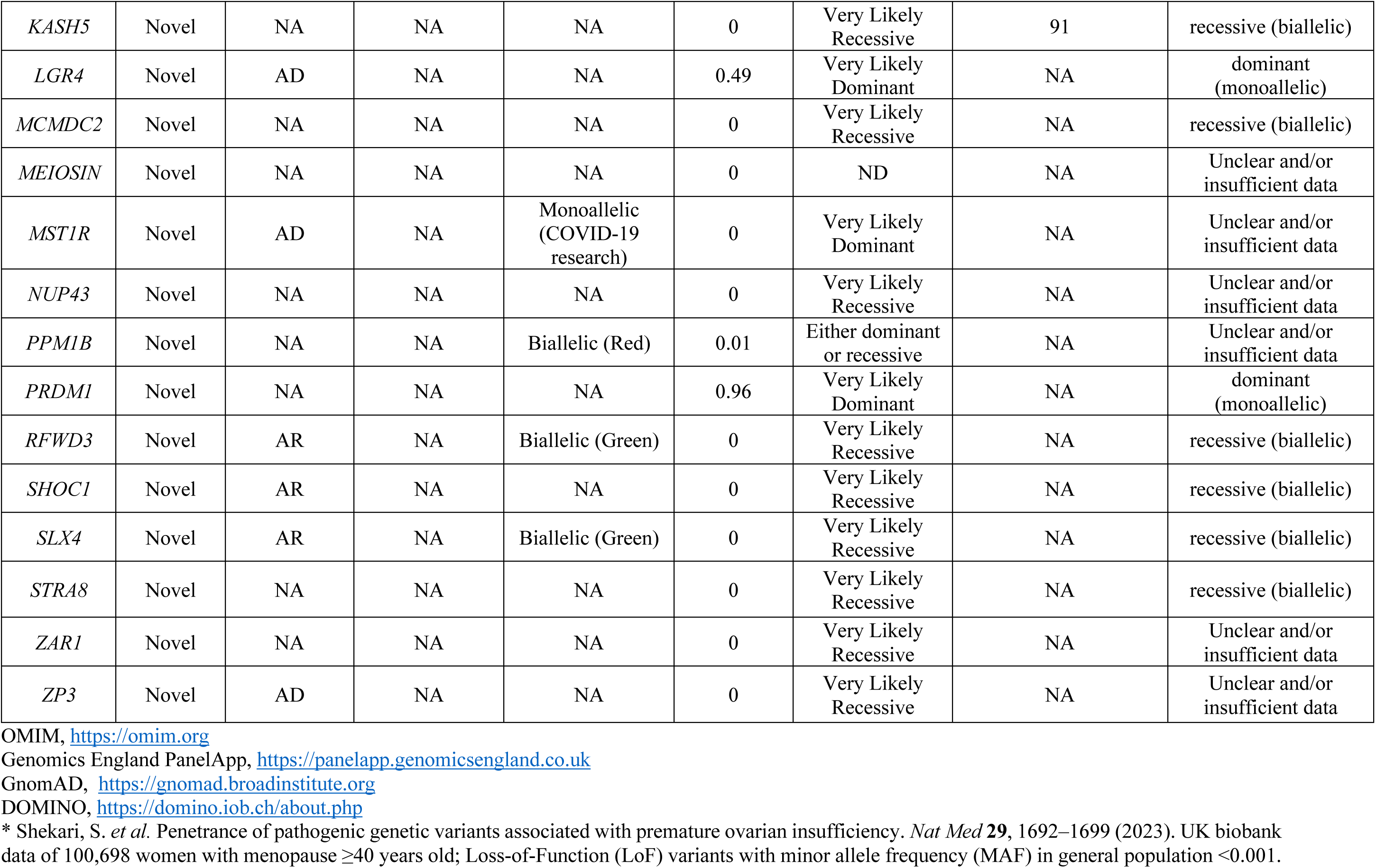
Assessment of inheritance mode of 79 genes reported by Ke et al with likely pathogenic or pathogenic variants in 242 Chinese patient with premature ovarian insufficiency (POI).

**Supplementary Table 2.**
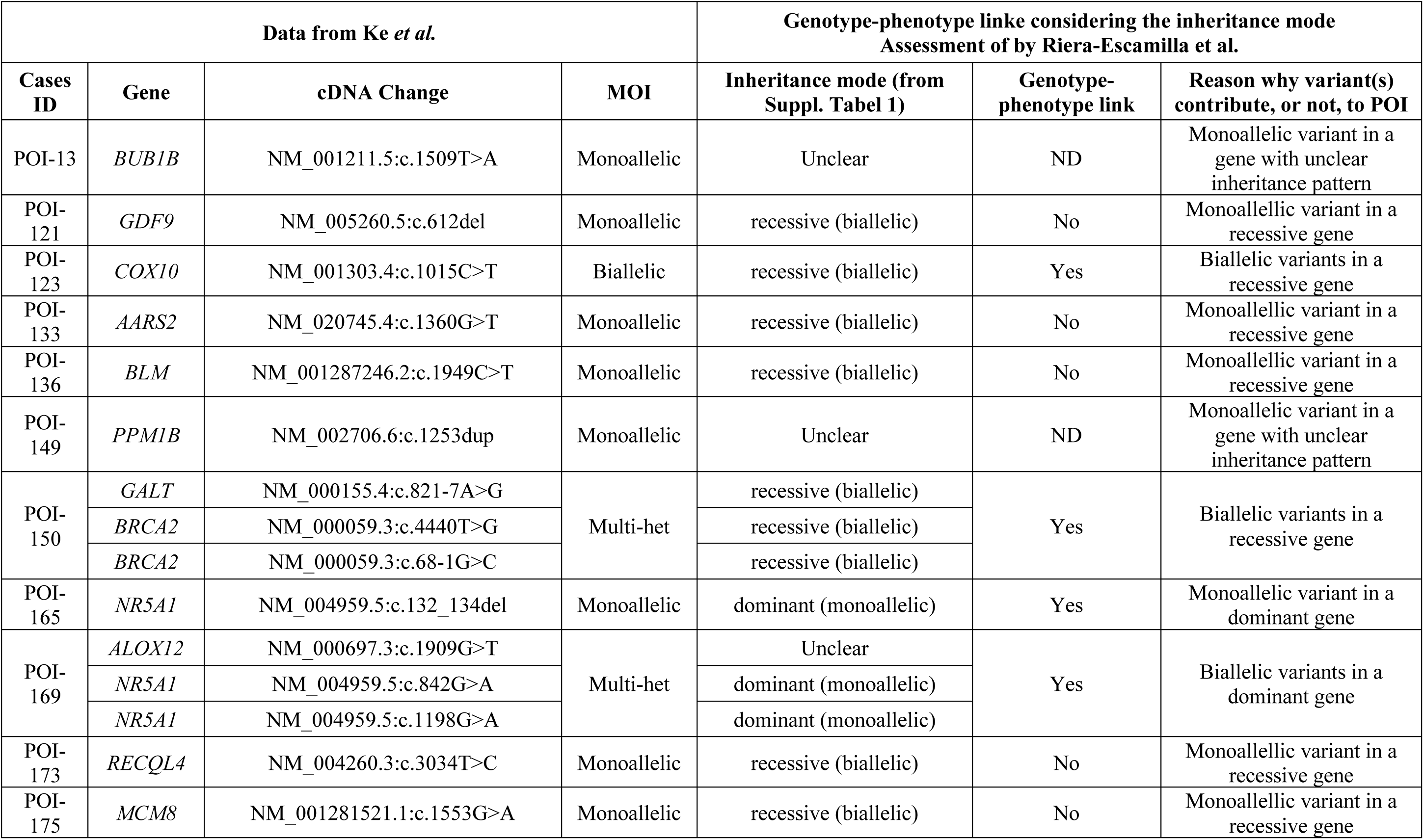
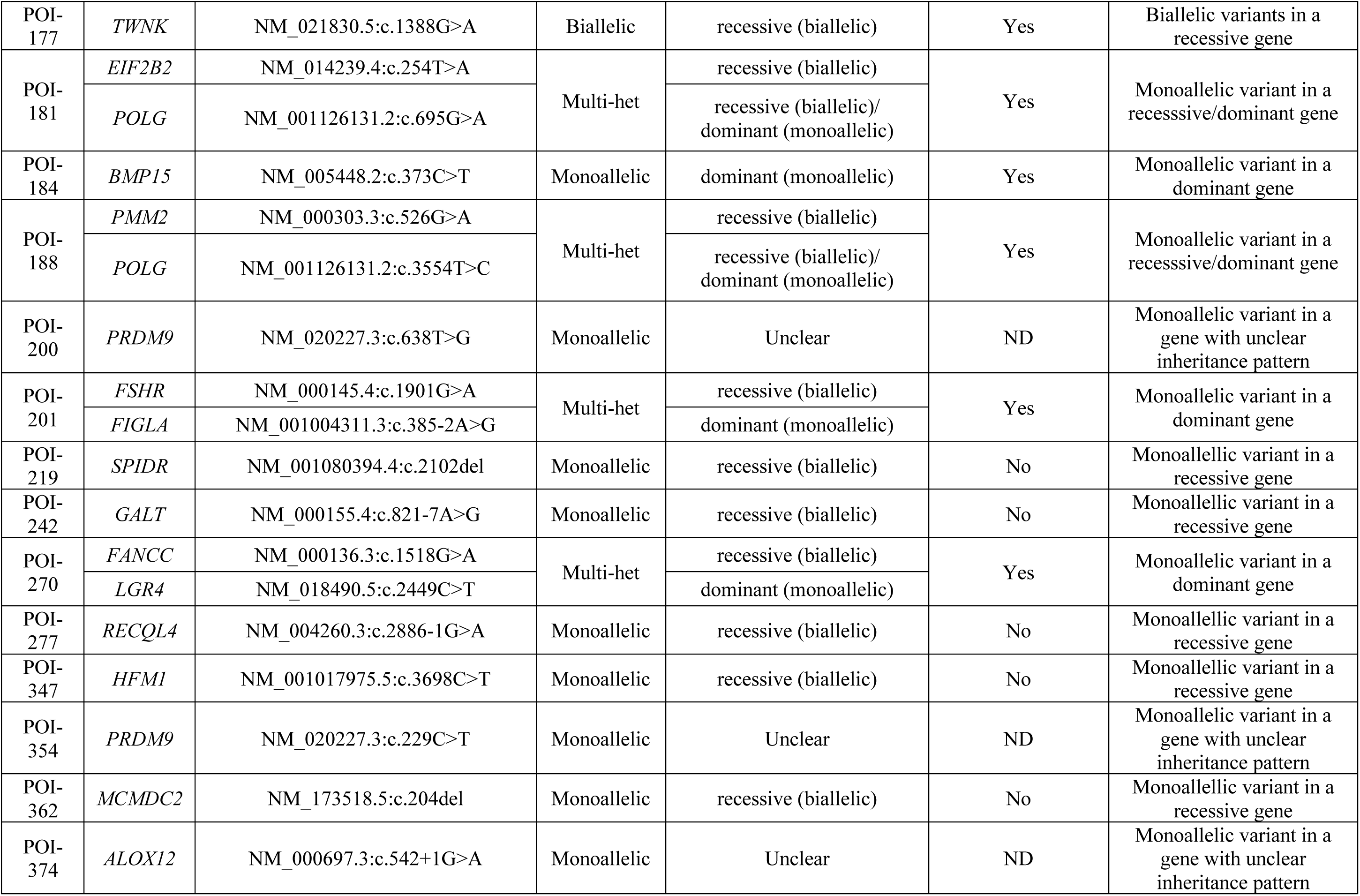
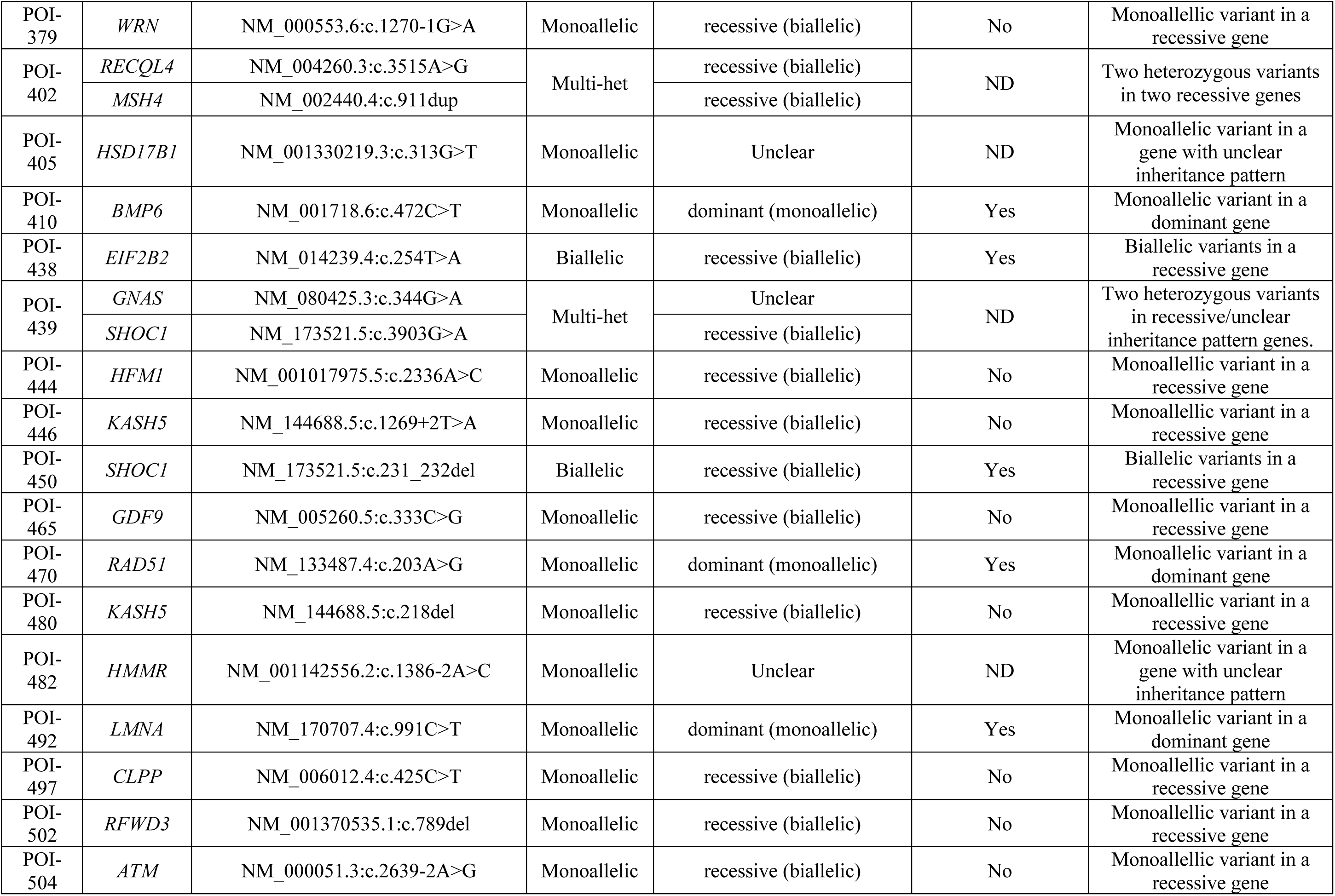
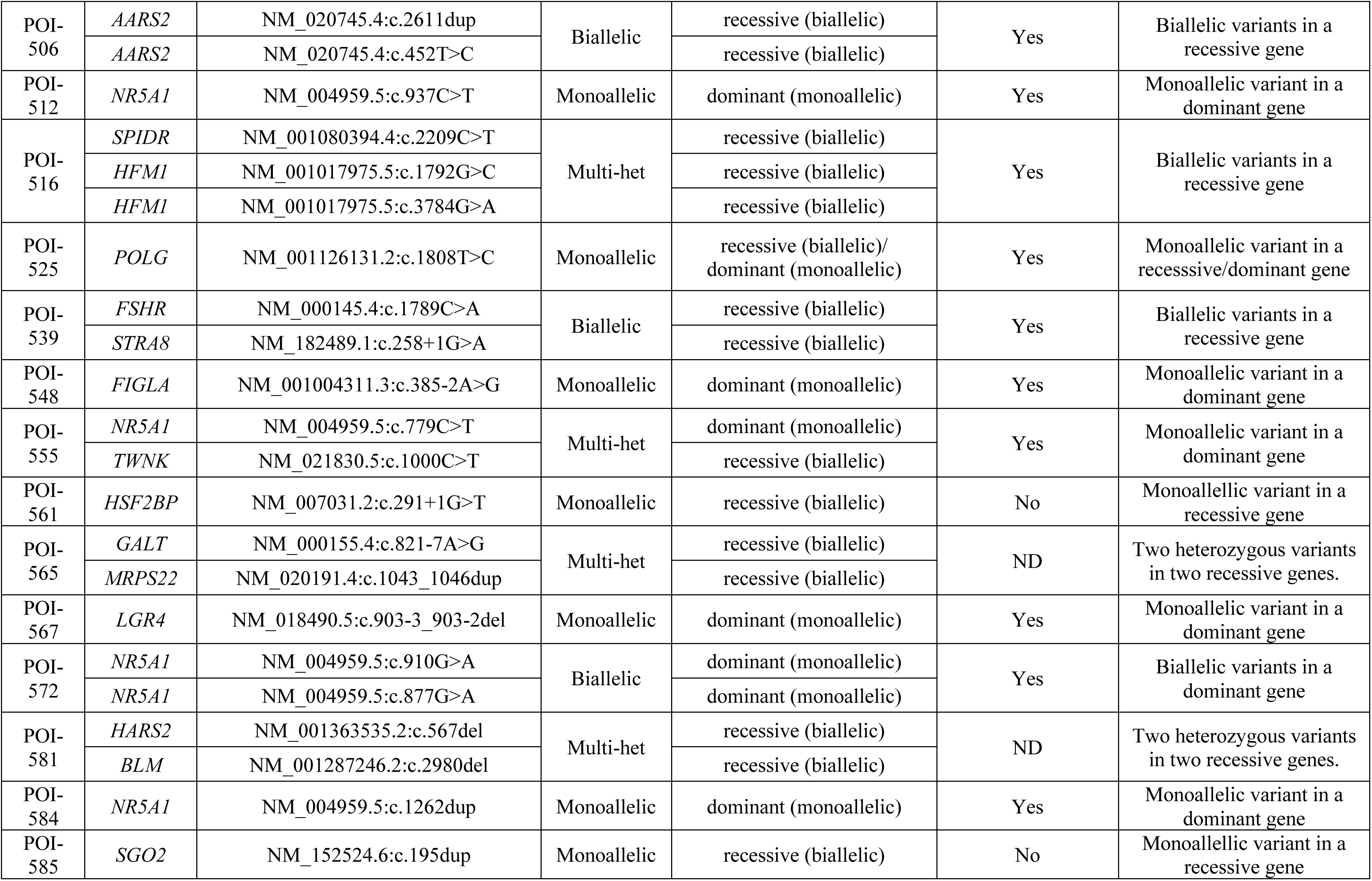
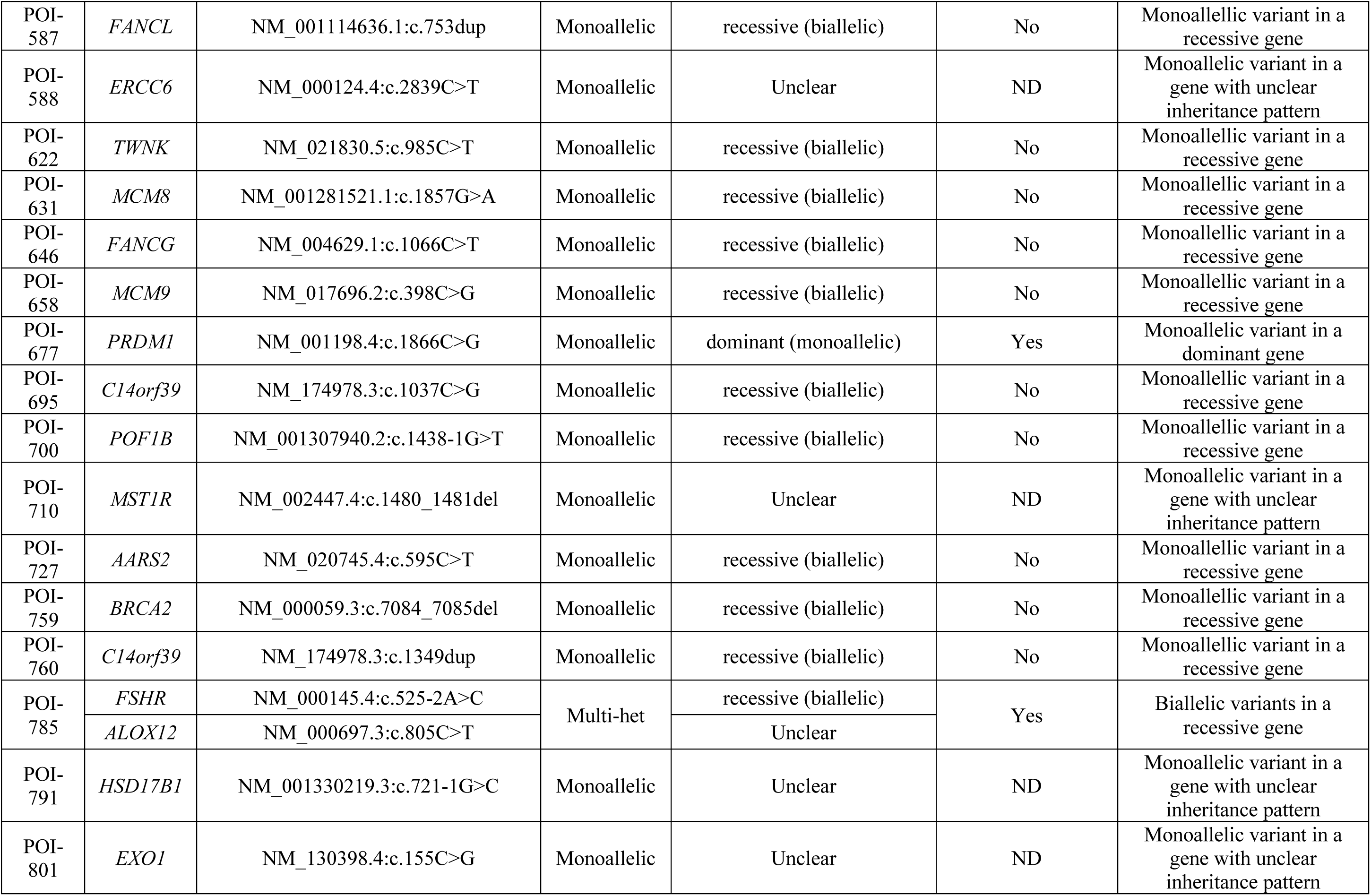
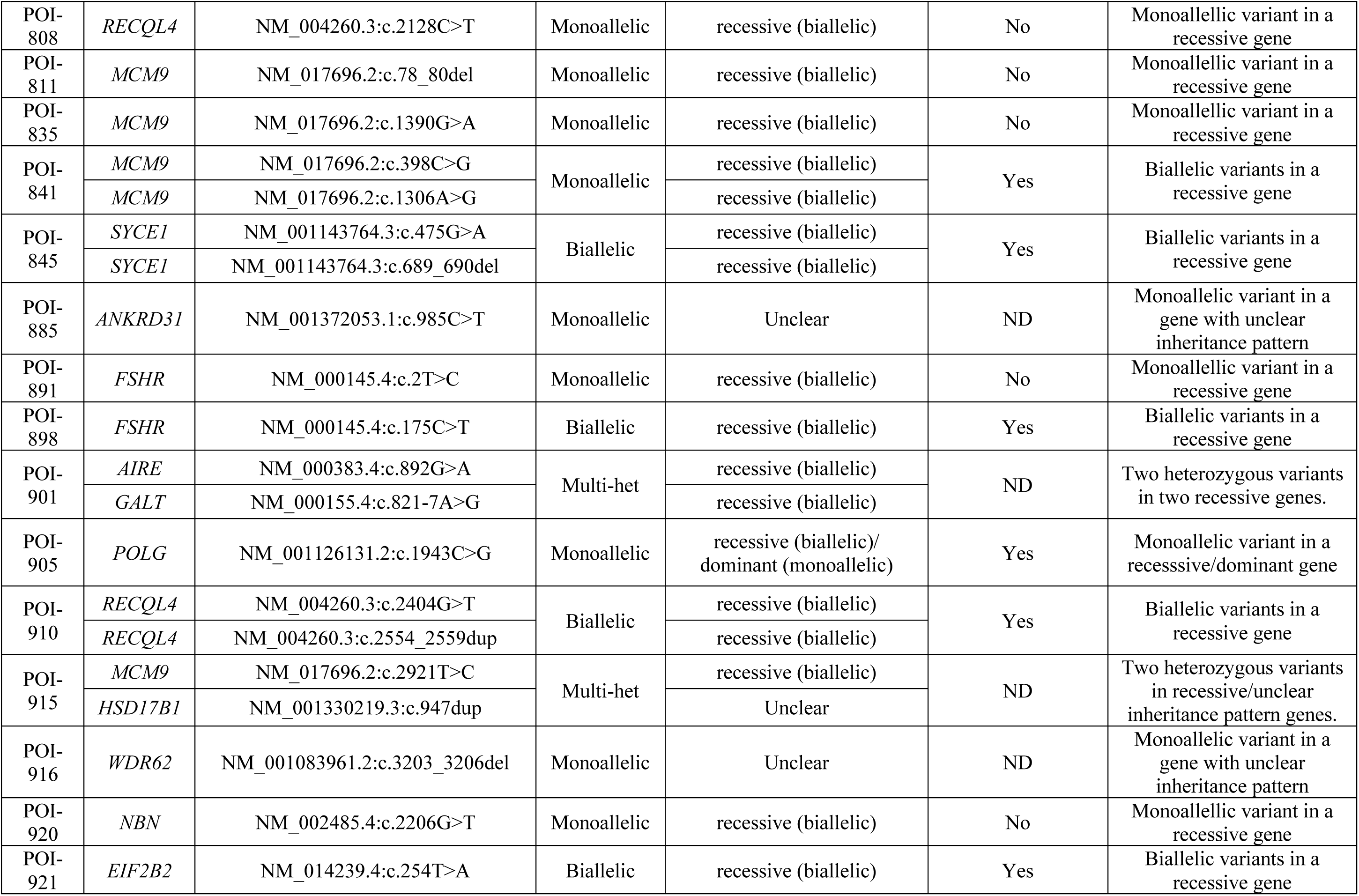
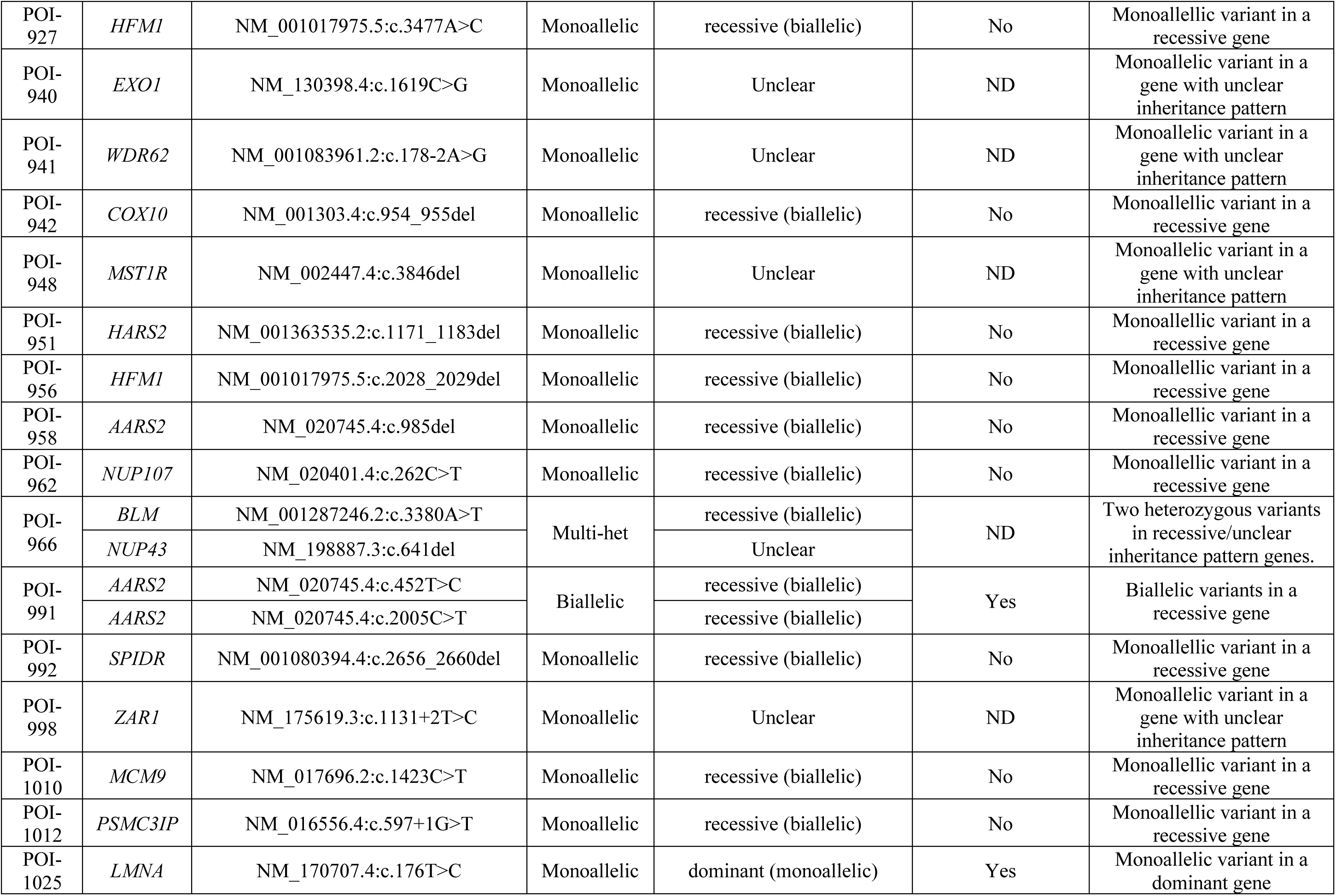
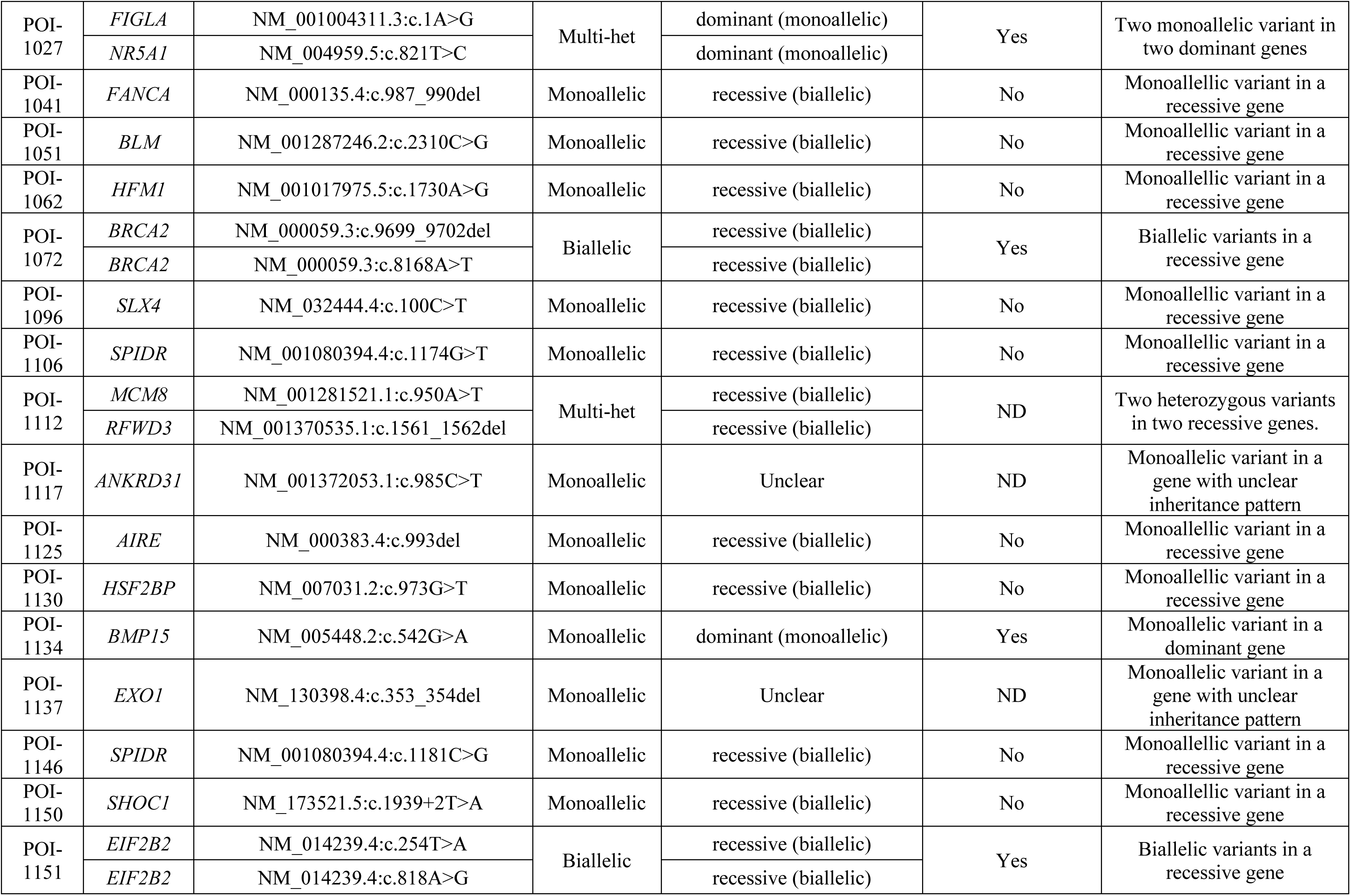
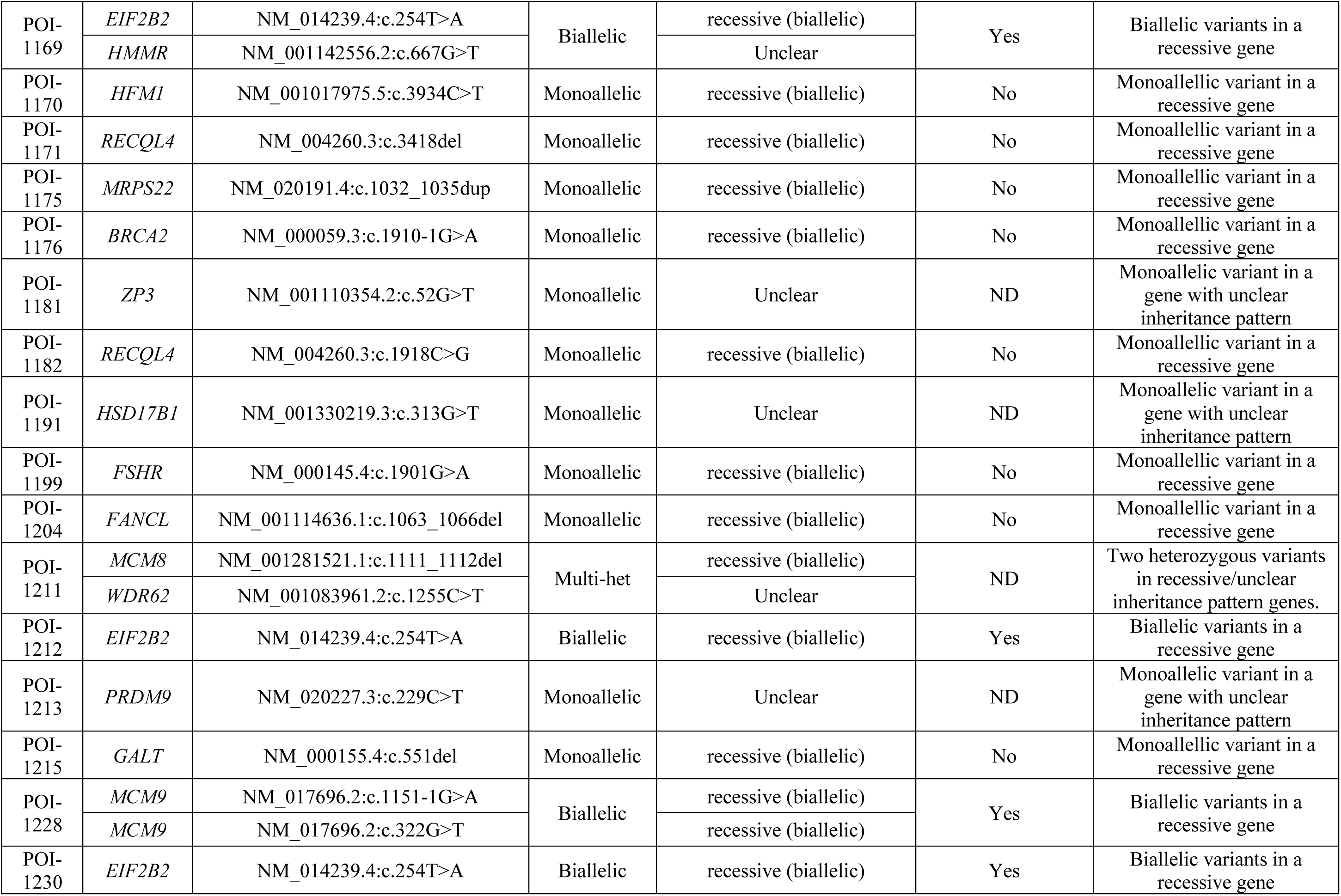
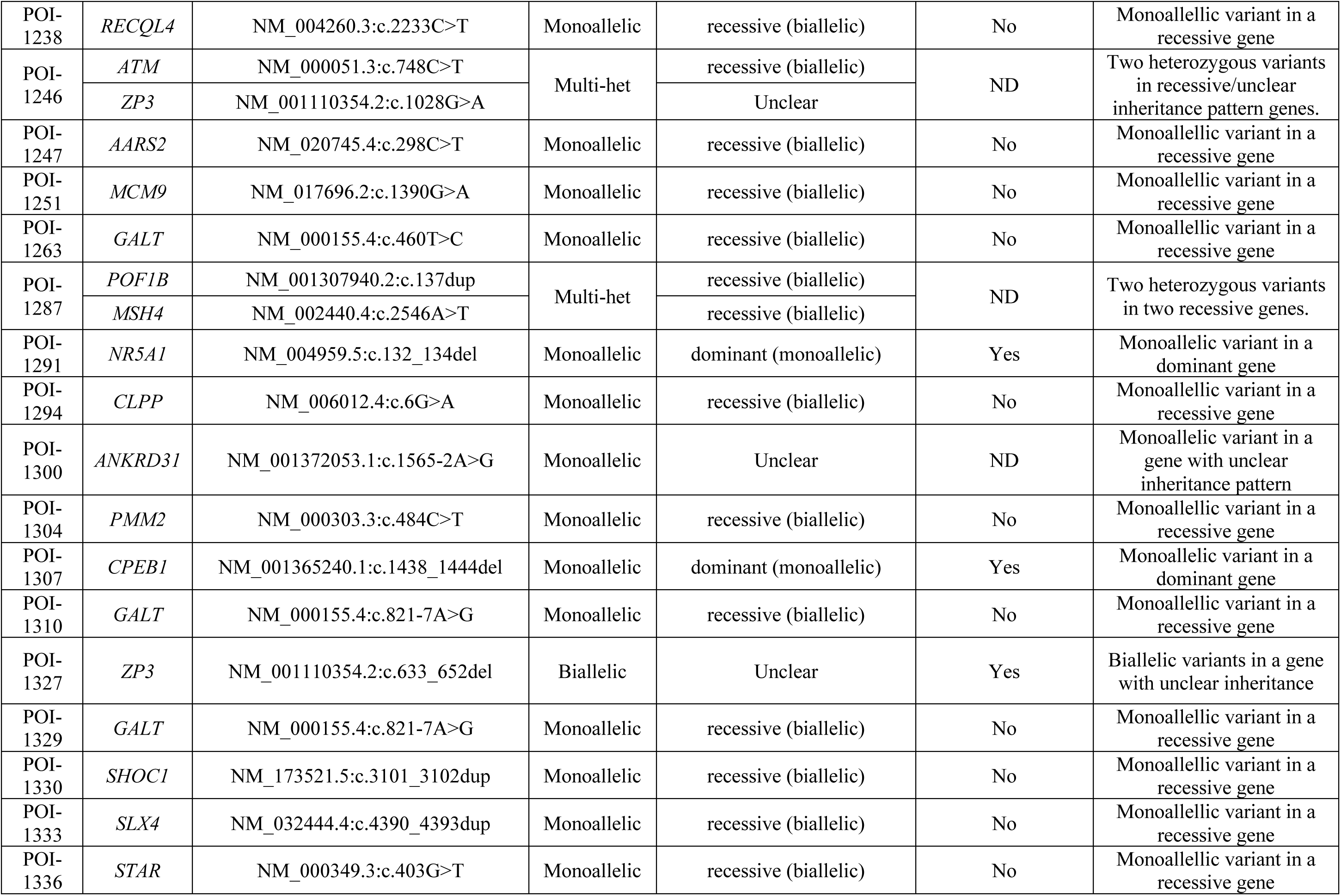
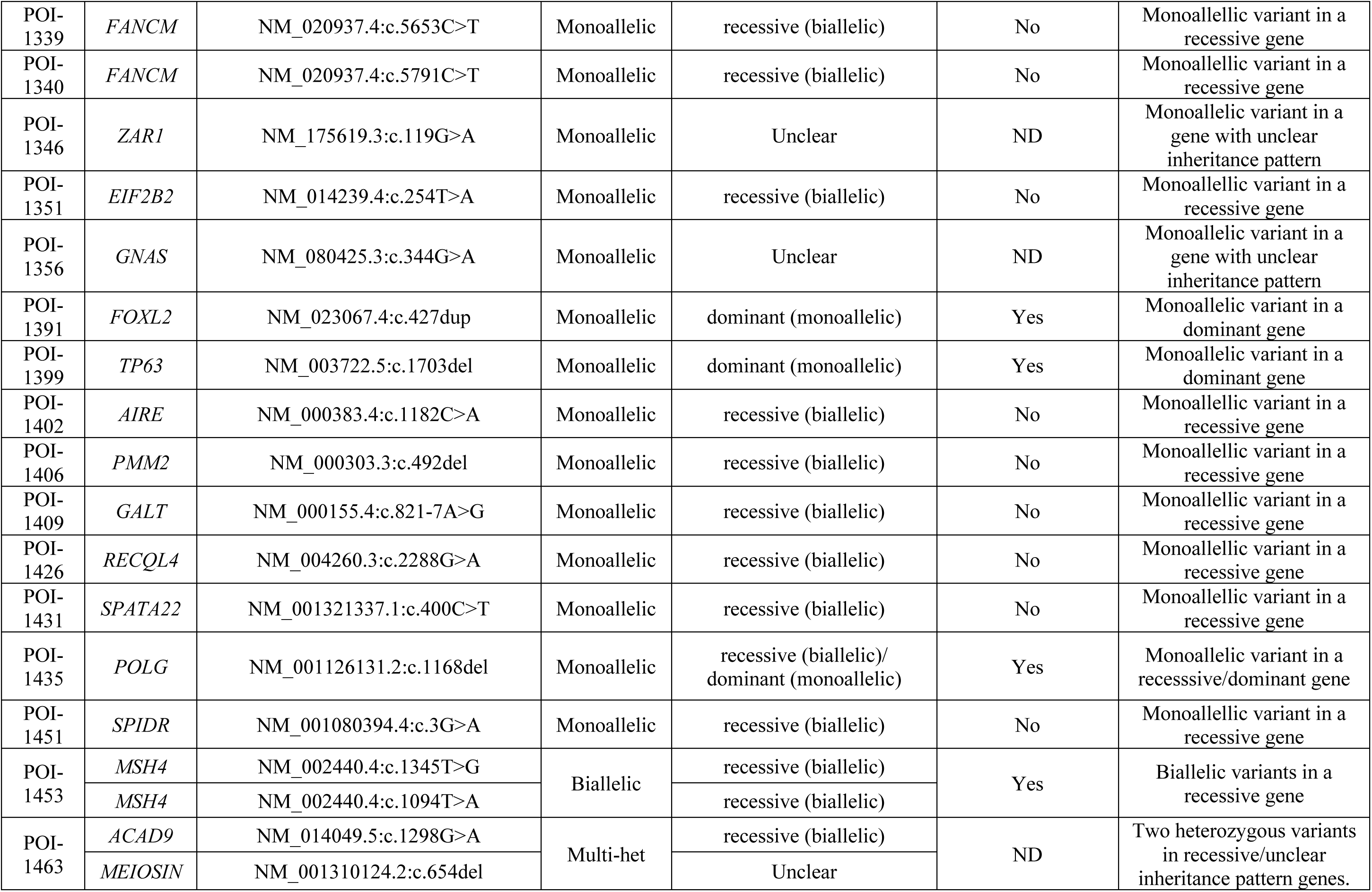
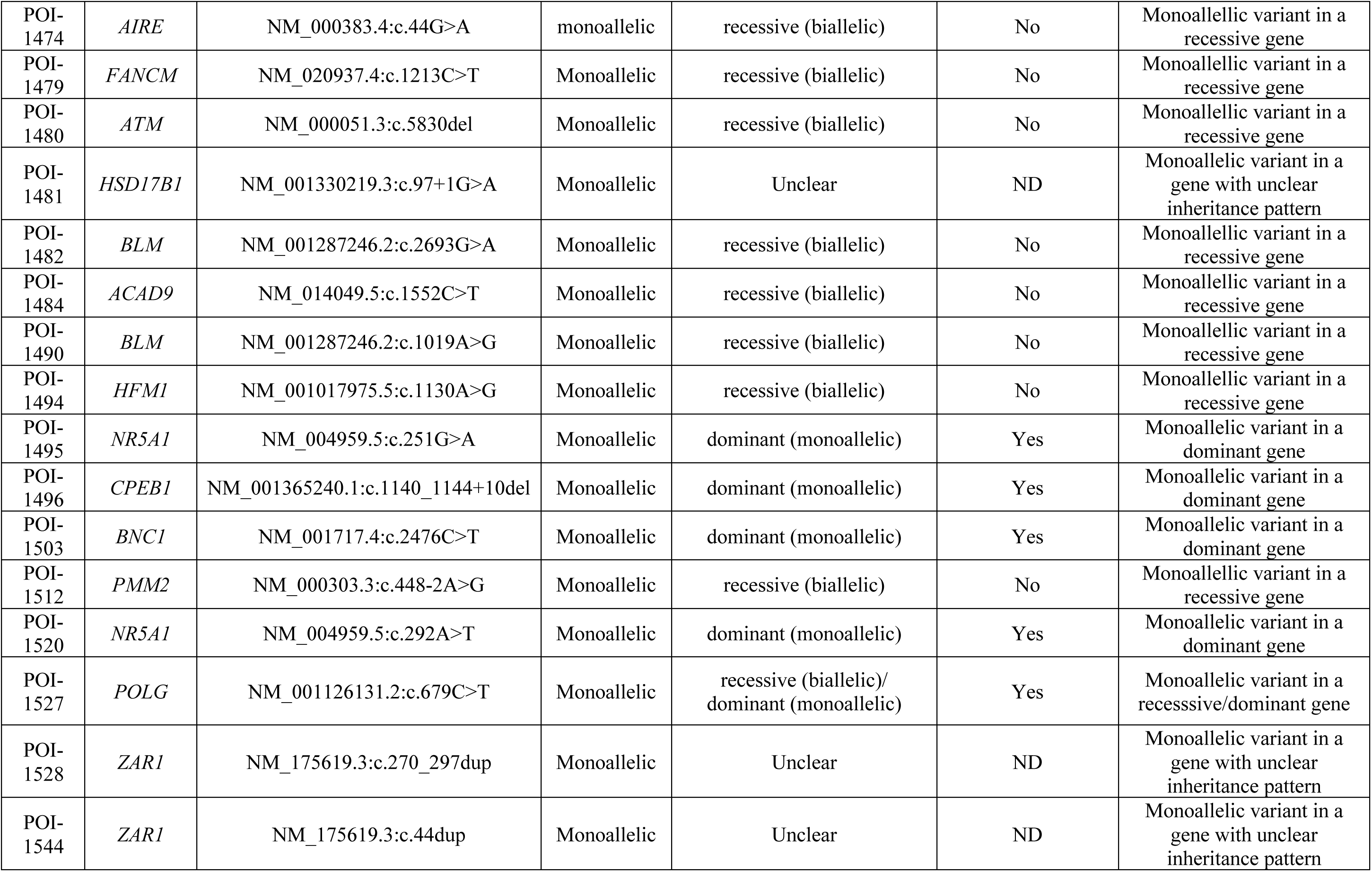
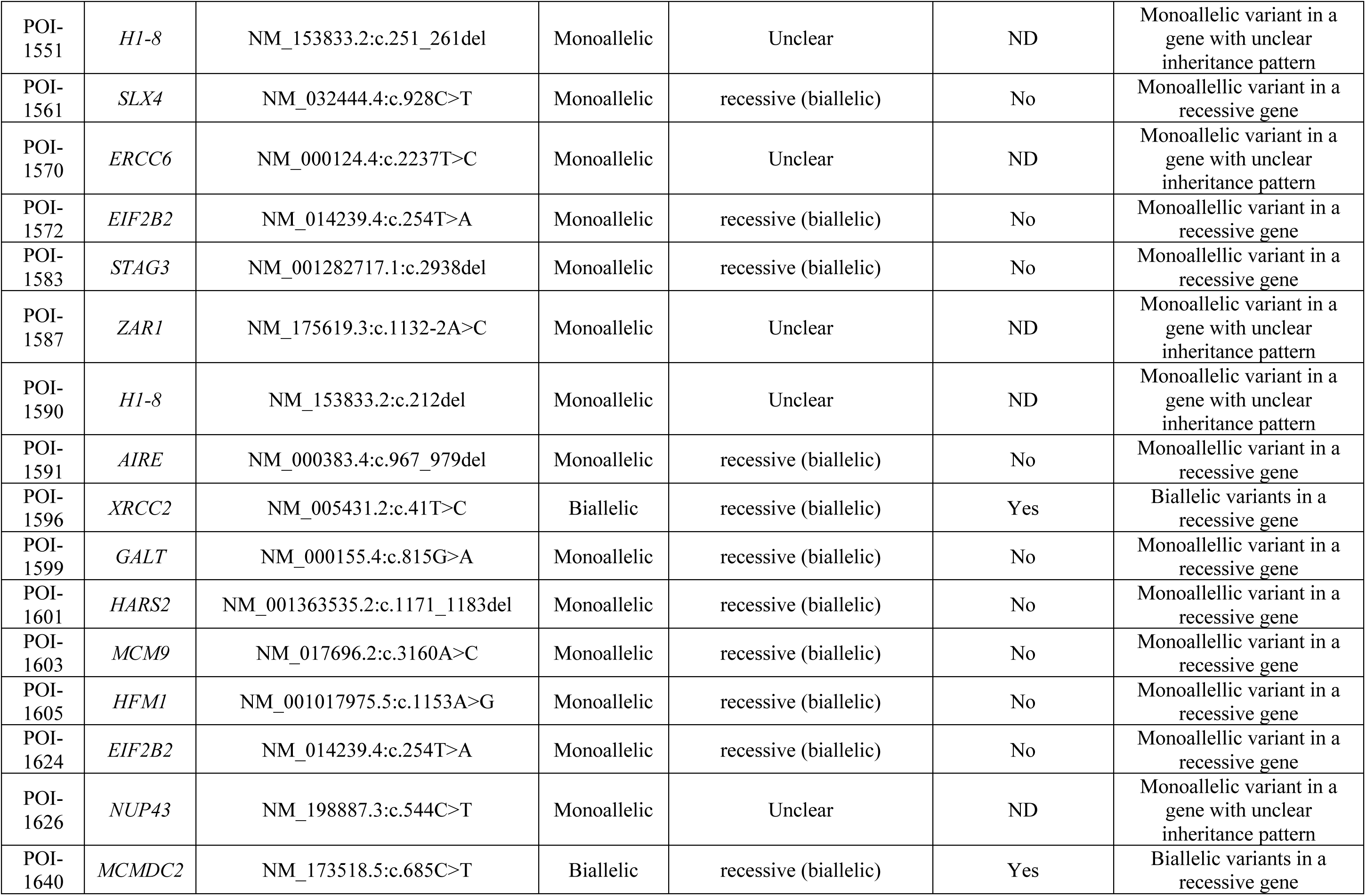
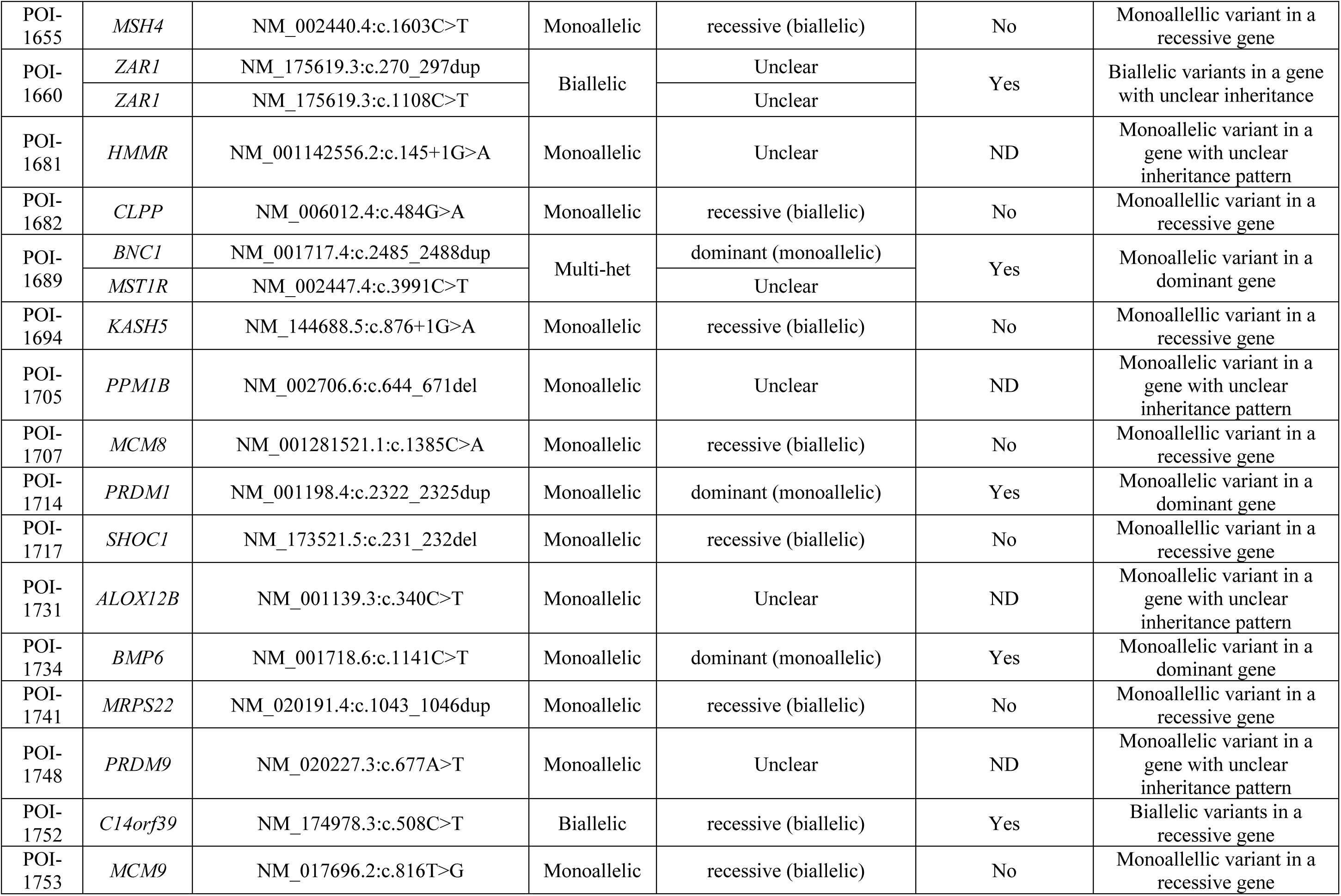
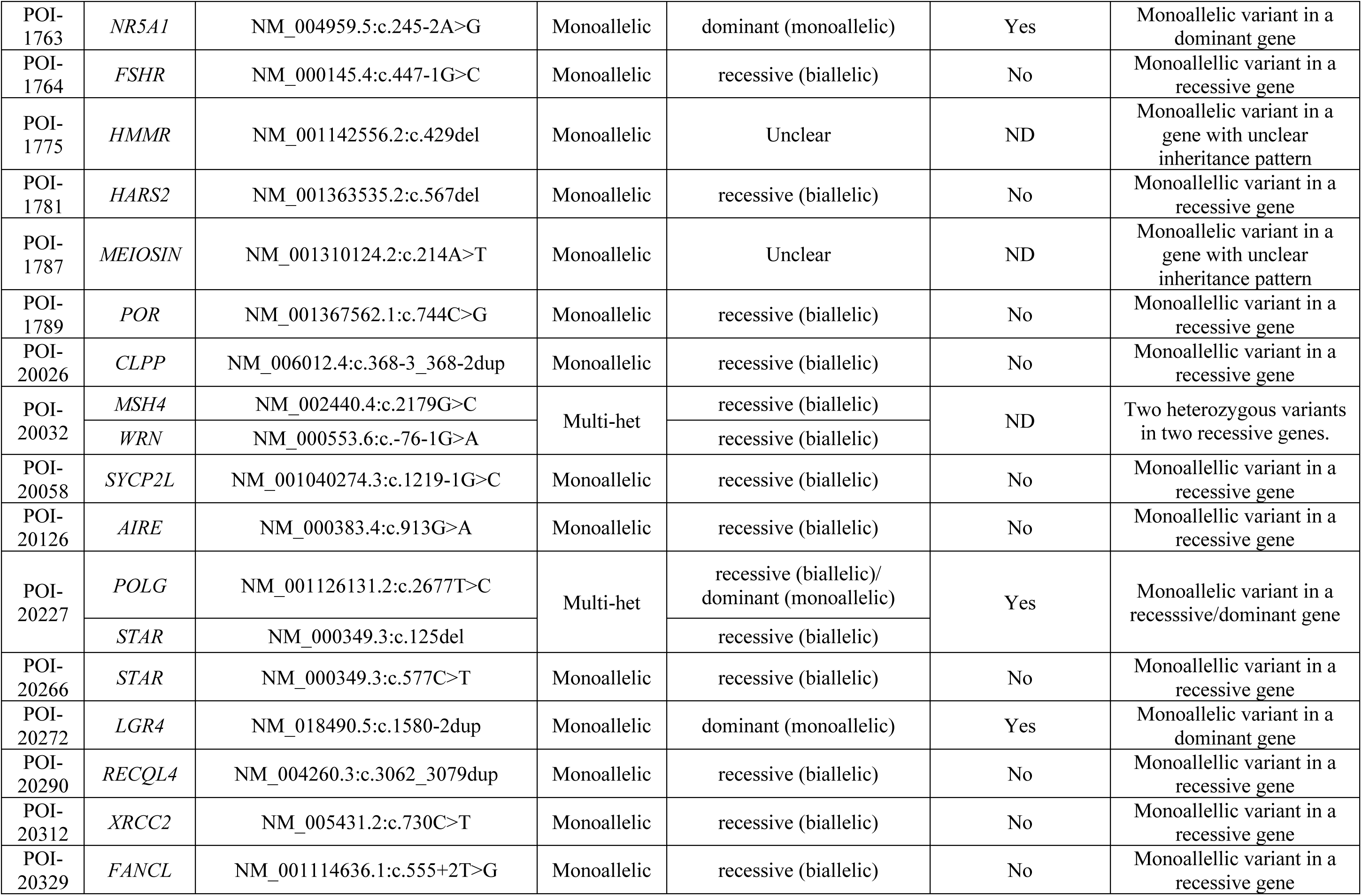
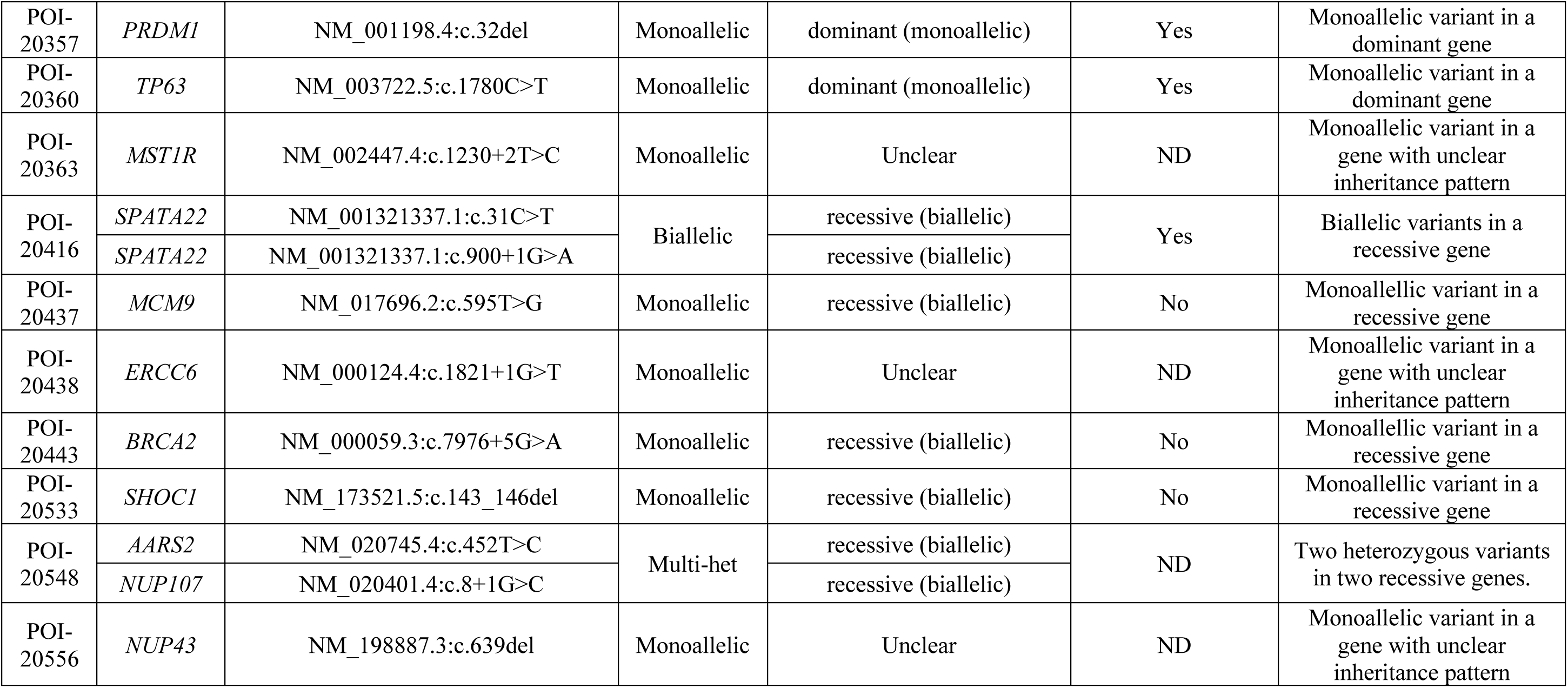
Assessment of reported genotypes in the context of known inheritance mode of monogenic conditions linked to the involved gene.

